# Repurposed medicines: a scan of the non-commercial clinical research landscape

**DOI:** 10.1101/2024.08.07.24310416

**Authors:** Sola Akinbolade, Ross Fairbairn, Alex Inskip, Rhiannon Potter, Aoife Oliver, Dawn Craig

## Abstract

Medicine repurposing is a strategy to identify new uses for existing medicines for the purpose of addressing areas of unmet medical need. This paper aims to provide horizon scanning intelligence on repurposed medicines that are evaluated by non-commercial organisations such as academia, and highlights opportunities for further research to improve patient health outcomes. A scan of the clinical landscape of non-commercially sponsored repurposed medicines is routinely conducted by the NIHR Innovation Observatory (IO). This ongoing project involves a horizon scan of clinical trial registries and the internal horizon scanning Medicines Innovation Database (MInD) of the NIHR IO to identify potential candidate medicines used as monotherapy or in combination to treat new indications outside the scope of their licensed indication. In addition to making these data publicly available, the output also supports the NHS England Medicines Repurposing Programme (MRP). The snapshot scan reported here (trials completing April 2020 to March 2023) identified a total of 528 technologies (meaning, a single product or combination of medicinal products targeting a specific indication in one or more related trials). The technologies were classified according to their characteristics and targeted therapeutic indications as well as revealing the least treated disease conditions. The candidate medicines identified in this scan could potentially receive tailored support towards adoption into practice and policy. The NIHR IO regularly provides this scan as a source of intelligence on repurposed medicines. This provides valuable insights into innovation trends, gaps, and areas of unmet clinical need.

## Introduction

Drug discovery and development require an adequate understanding of the intended disease condition, identification of potential candidate molecules, established methods for drug production and the initiation of clinical trials to test the efficacy, effectiveness, and safety of the drug.^1^ The journey of new drug discovery and the overarching target to address unmet clinical needs imposes major challenges to the pharmaceutical industry.^2,3^ The potential for serious unacceptable adverse effects or inadequate efficacy profiles, coupled with long timelines and high cost implications limit the successful development of new active substances.^4,5^ If substantial reductions in the process, associated costs and time for drug development could be realised, this would provide clinicians and patients with early access to effective medicines.

Medicine repurposing, also known as drug repositioning, can be defined as a strategic process of identifying new uses or indications for approved medicines beyond their original indication.^3^ The repurposing strategy serves as an opportunity to address areas of unmet treatment need in a timely manner while reducing costs and associated risks of novel drug development.^6^ For example, the urgent need for effective treatments for COVID-19 required the early discovery of suitable medicines that showed great promise to treat the disease, and this led scientists to use repurposing strategies.^7^ Many years of research have discovered the potential for existing medicines to treat conditions outside their licensed indications.^8^ In the 1980s, the analgesic aspirin was repurposed as an antiplatelet aggregation drug for cardiovascular disorders, and more recently has been investigated for cancer prevention. Sildenafil, an investigative drug for the treatment of coronary artery disease, hypertension, and angina pectoris, produced unexpected side effects which led to its repurposed indication for the treatment of erectile dysfunction.^8,9^

The concept of medicine repurposing has been pursued not only by the pharmaceutical industry, but also by research institutes and academia.^10^ Academic and investigator-initiated research projects are usually focussed on the achievement of a scientific breakthrough that could improve patients’ health and attract funding or partnerships, rather than a commercial gain or the accomplishment of a strategic business model.^10^ However, by exploring repurposing without industry collaborations, academic researchers typically face a number of financial, infrastructural, operational, and regulatory hurdles.^11,12^ The lack of access to preclinical and clinical data on discontinued/failed candidate molecules or drugs protected by patents held by pharmaceutical companies, restricts the exploration of potential candidate molecules to generic (off-patent) drugs.^10^ Further, the financial burden of conducting investigator-led studies has been identified as one of the major barriers to repurposing.^12^ It is worth noting that, whilst a pharmaceutical company (as a licence holder for an approved medicine) would only need to submit a clinical variation to include a new indication, a non-commercial organisation (i.e. academic researchers) would be required to follow a lengthy full application process to obtain approval or marketing authorisation for a candidate medicine, and may not be able to fulfil the legal, financial and regulatory responsibilities of a licence holder.^13^ These issues and requirements can be challenging to academic researchers, which in turn may hinder the development of the repurposed medicine or stop the exploration in the first instance.

The establishment of collaborations between pharmaceutical companies and academic researchers appears to offer an opportunity to overcome many of these hurdles.^3,10^ These partnerships could provide a platform to combine expertise in drug research with experience in commercial drug development.^10^ The initiation of government-sponsored schemes could also provide support and promote repurposing opportunities for non-commercial organisations in terms of providing financial incentives, supporting evidence generation and adoption into the healthcare system.^13^

Despite the hurdles outlined, there are many clinical trials being conducted and sponsored by non-commercial organisations hoping to enable access to new treatment options for the benefit of public health. Given the significant unmet need in some disease areas, there is an increased focus on medicines repurposed to meet these needs; this activity is mostly driven by academia or research institutes.^12^ However, there is a growing pool of candidate medicines which are not being pursued for repurposing by pharmaceutical companies either due to failed efficacy in previous studies or irrelevance to their business model.^3^ These potential candidate medicines could provide opportunities for academia to explore and discover new indications to which the medicine may be used.

The aim of this paper is to provide horizon scanning data and intelligence on repurposed medicines in clinical development by non-commercial organisations; and offer valuable insights for future medicine repurposing research as well as potential tailored support towards their adoption into practice and policy to improve patient health outcomes.

## Method

In 2022, the National Institute for Health and Care Research (NIHR) Innovation Observatory (IO) began an ongoing horizon scanning project in collaboration with the NHS England Medicines Repurposing Programme (MRP) to identify repurposed medicines in clinical development by non-commercial organisations.^13^ The project involves the horizon scanning of clinical trial registries (ClinicalTrials.gov and EU Clinical Trials Register) and the IO’s internal horizon scanning Medicines Innovation Database (MInD) to identify potential candidate medicines used as monotherapy or in combination to treat new indications outside the scope of their approved or licensed indication.^14^

A detailed search strategy was developed to identify potential repurposed candidates in clinical trials using the following search terms: *“interventional”* for study type selection; *“Not yet recruiting OR Recruiting OR Active, not recruiting OR Completed OR Enrolling by invitation OR Suspended OR Unknown”* for study status; *“All Others (individuals, universities and organisations)”* for funder type; and primary completion date *“01/04/2020 – 31/03/2023”*. The identified trials were uploaded to an internal IO filtering tool to remove clinical trials that do not have at least one location in USA, European Union, United Kingdom, Australia, or Canada. Additional trials were identified from IO’s MInD based on the appropriate primary completion date range and ‘repurposed technology’ tag. The IO’s MInD is an internally facing horizon scanning database that is populated with data from clinical trial registries, direct engagement with companies, news media, and <u>UK PharmaScan</u>. The screening of these data sources utilised a combination of traditional scanning methods (manual), automated and novel AI/machine learning techniques. A clinical trial was identified as ‘repurposed’ based on the current UK Marketing Authorisation status of the experimental medicine, which was manually searched from <u>Electronic Medicines Compendium (EMC)</u> or <u>Medicines and Healthcare products Regulatory Agency (MHRA)</u>. Additionally, clinical trials were excluded based on these criteria: complementary and alternative medicines (CAM), herbal medicines, traditional Chinese medicines (TCM), dietary supplements, procedural or surgical interventions, and medicines repurposed to treat COVID-19.

An initial pilot scan captured a large dataset of repurposed candidate medicines from Phase I/II up to III. However, following the completion of this pilot, the selection criteria were further modified, and changes were applied to the search strategy for future scans; this involved excluding Phase I/II and II trials for non-rare conditions. This modification provided focus on promising repurposed medicines targeting rare conditions in Phase I/II to III, and non-rare conditions in Phase II/III and III.^14^ The data provided in this paper are from the repurposed medicines scans conducted for clinical trials with primary completion dates from 1 April 2020 – 31 March 2023, and are limited by the specified inclusion criteria for this project (Table 1).

**Table 1.** Characteristics for inclusion of clinical trials and their investigative medicines in the repurposed medicines scan.

| Criteria | Characteristics for inclusion |
| --- | --- |
| Medicinal product | Marketing Authorisation in the UK |
|  | Off-patent (generic) repurposed medicines |
|  | On-patent (branded) repurposed medicines |
|  | Reformulated generic medicinal products |
|  | Biosimilars used outside the Marketing Authorisation of the originator product |
|  | Monotherapy or combination therapy |
| Clinical trial phase | Phase I/II, II, II/III and III for rare disease conditions |
|  | Phase II/III and III for non-rare disease conditions |
| Trial locations | UK, EU, USA, Australia, or Canada |
| Trial sponsorship | Non-commercial sponsors such as academia, hospitals, charities |
|  | Non-commercial sponsors with industry collaborators |
| Trial primary completion dates (PCDs) | 1 April 2020 – 31 March 2023 (Dates not applicable to trials sourced from the EU Clinical Trials Register) |

## Results

The repurposed medicines scan identified a total of 528 eligible technologies sourced from clinical trial registries and the IO’s MInD. ‘Technology’ refers to a single or combination of medicinal products targeting a specific indication in one or more related trials. For technologies with more than one clinical trial, priority was given to the trial with highest trial phase, and which had a primary completion date within the specified range (as the ‘main’ trial).

### Clinical trial characteristics

Analysis of the 528 technologies showed 93 technologies (17.6%) were no longer in development (terminated, suspended, withdrawn, prematurely ended or unknown status); 142 technologies (26.9%) had completed, and 293 (55.5%) were in active clinical development (Figure 1).

**Figure 1.**
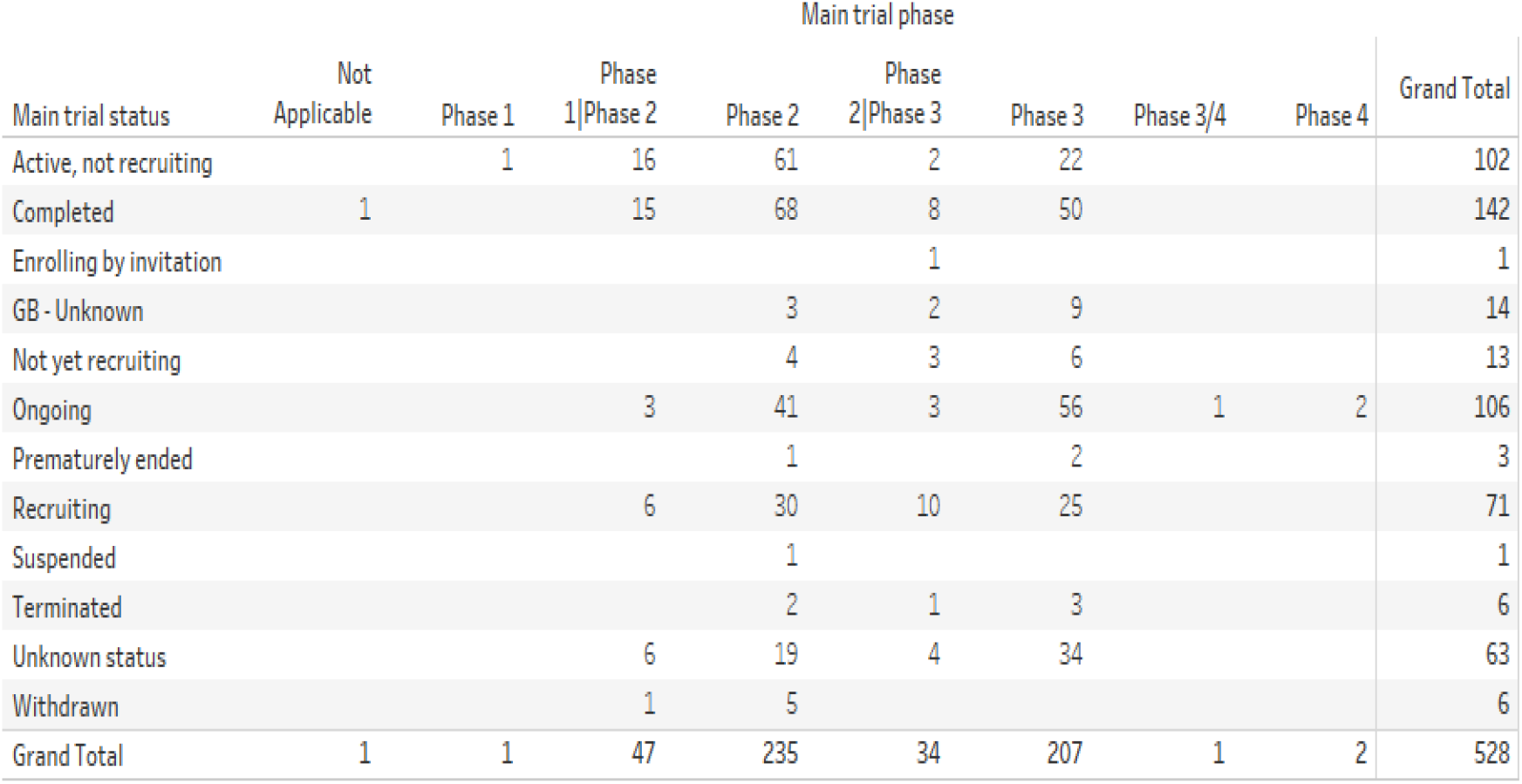
Overview of the clinical trial phases and status of the technologies.

There was a noticeable increase in the number of eligible clinical trials that started between 2017 and late 2020, with a total of 252 trials during this period. Earlier than 2010, there were about six eligible trials evaluating repurposed indications according to the inclusion criteria (Figure 2).

**Figure 2.**
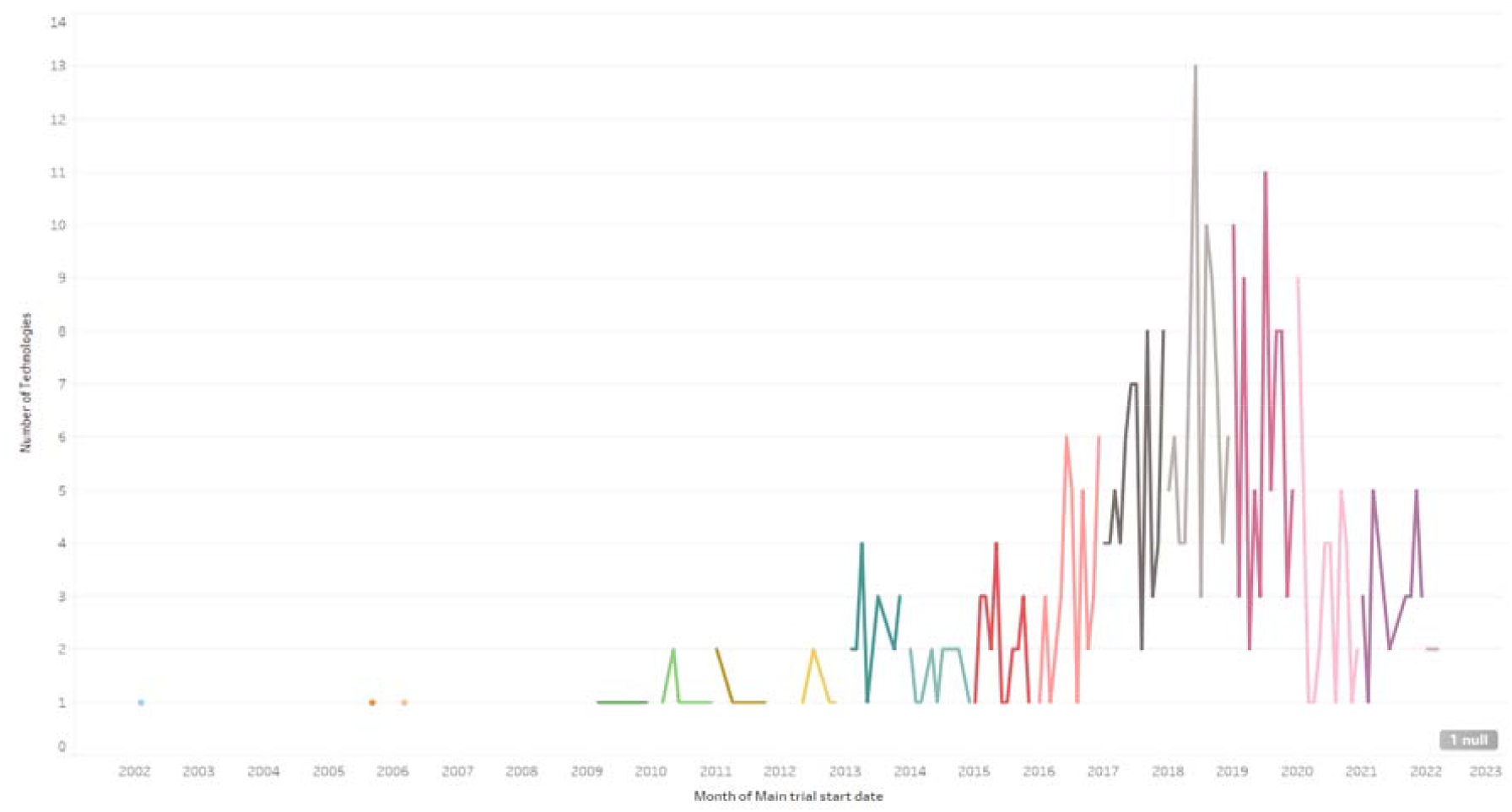
Distribution of trial start dates for the technologies

### Classification of the repurposed technologies

The identified repurposed technologies were categorised using the IO’s classification system for innovative medicines, namely: repurposed technologies in combination; off-patent/generic repurposed technologies; and branded/on-patent repurposed technologies.^15^ There were 206 (39%) technologies evaluating the synergistic effect of combining repurposed medicines; while 322 (61%) tested medicinal products as monotherapy. The combination therapies were either on- or off-patent repurposed medicines, or both. Further categorisation of the 322 monotherapy technologies revealed that 182 (56.5%) tested the efficacy of off-patent/generic medicinal products in a new indication; while 140 (43.5%) evaluated medicinal products still covered under UK patent protection (on-patent/branded).

### Categorisation according to therapeutic areas and disease conditions

Further analysis of the 528 technologies showed the range of therapeutic areas explored, with the largest proportions investigating haematological cancers and lymphomas (21.4%), gastrointestinal cancers (10.2%), and neurological disorders (8%) (Figure 3). The least investigated therapeutic areas were male reproductive cancers (0.4%), urological cancers (0.2%), and women’s and men’s health disorders (not otherwise classified) (0.2%) (Figure 4).

**Figure 3.**
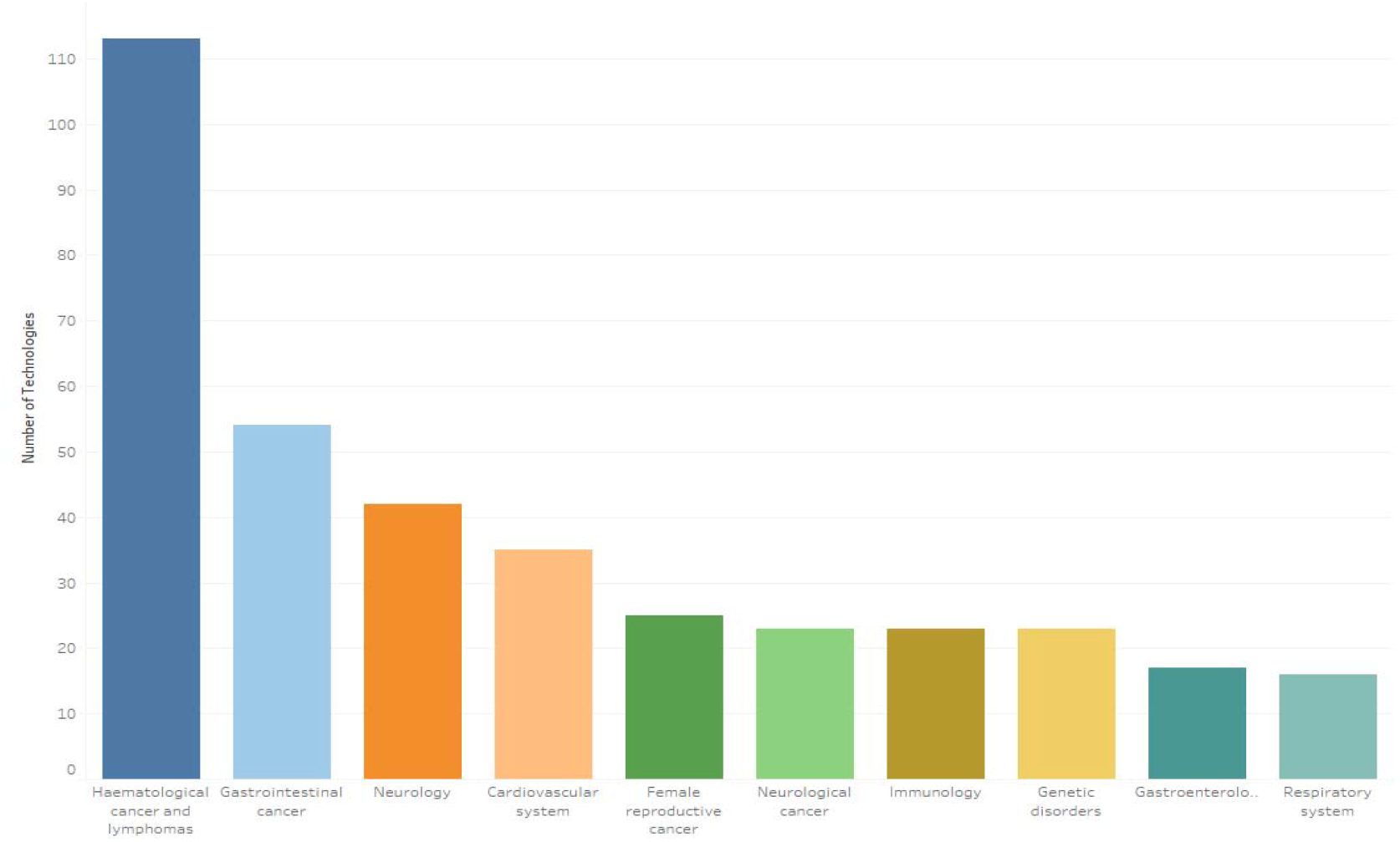
Top ten therapeutic areas investigated in the repurposed medicines scan

**Figure 4.**
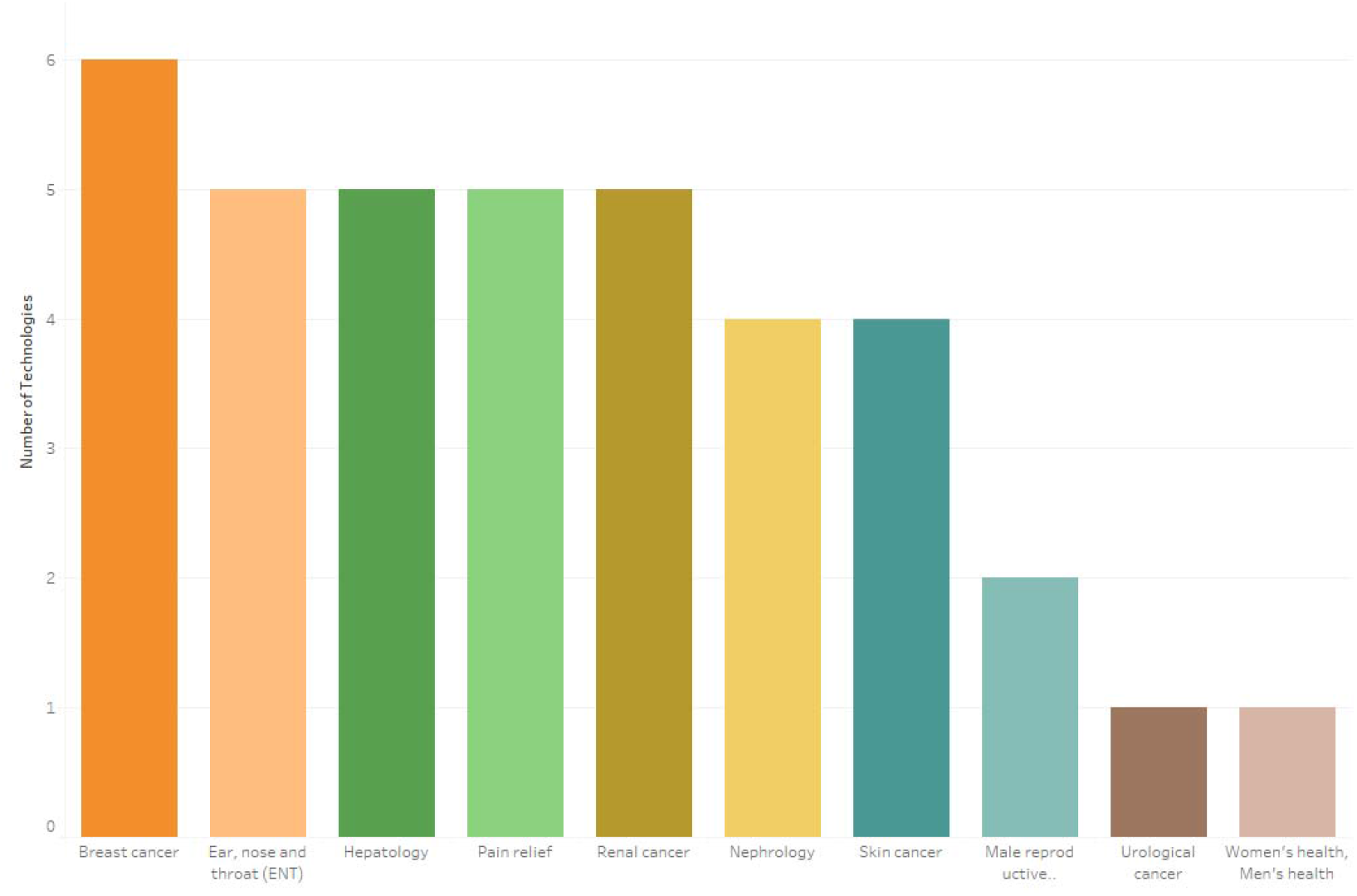
Bottom ten therapeutic areas investigated in the repurposed medicines scan

Across all the therapeutic areas identified, the technologies were evenly split between cancer (51.9%) and non-cancer (48.1%) indications.

The data revealed that 357 technologies (67.6%) targeted rare disease conditions, with multiple myeloma, pancreatic cancer, and glioma being the most treated conditions (Figure 5).

**Figure 5.**
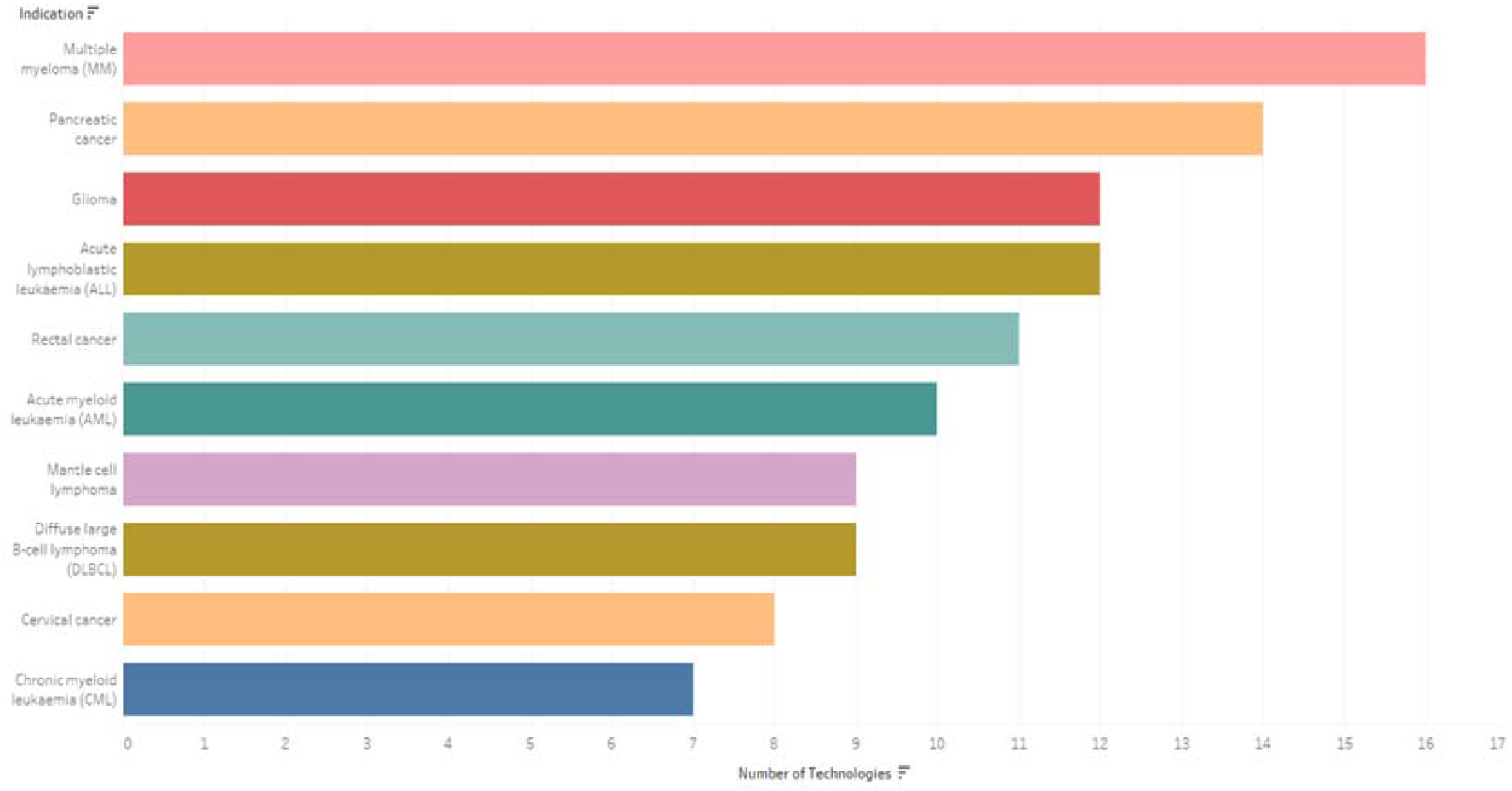
Top ten indications for rare disease conditions

Additionally, categorising the identified technologies according to their therapeutic classes revealed that monoclonal antibodies (30.1%), kinase inhibitors (19.7%) and corticosteroids (7.8%) were the most frequently repurposed classes (Figure 6).

**Figure 6.**
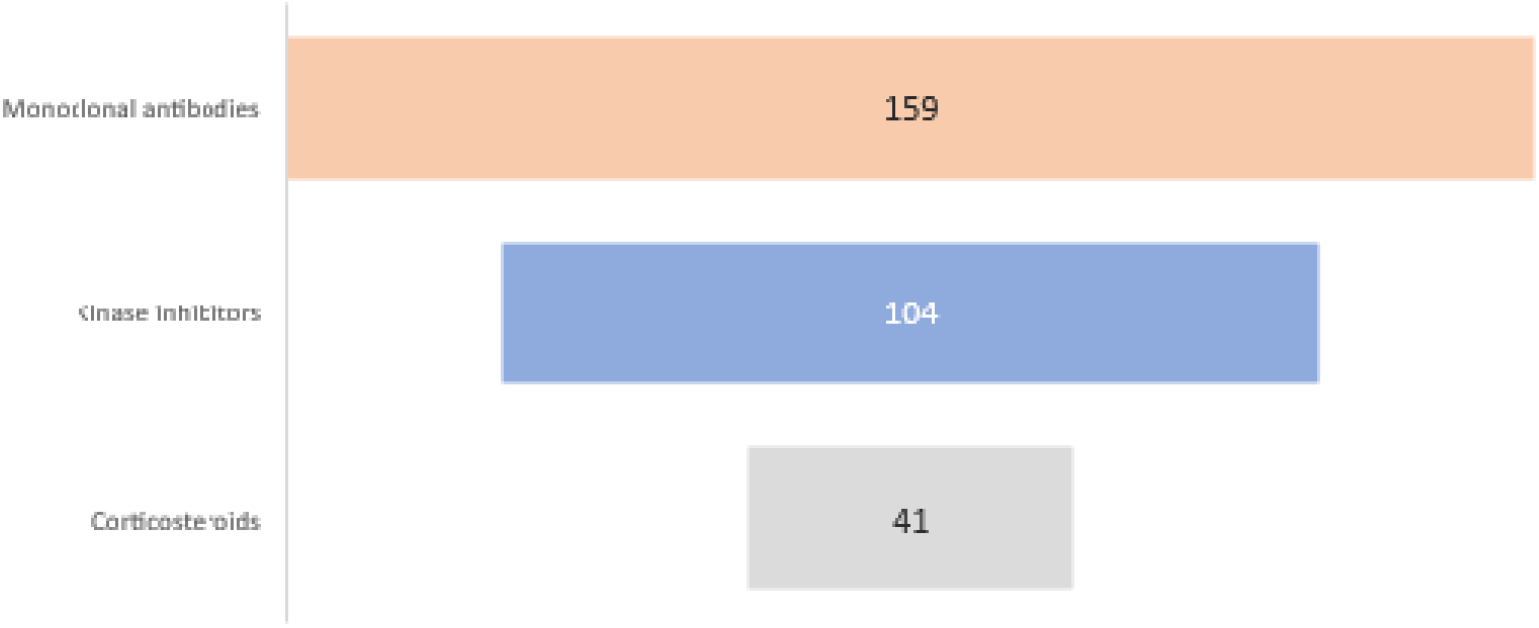
Number of technologies for the top three therapeutic classes identified. Each class may have been used in combination with themselves, other therapeutic classes, or as monotherapies.

## Discussion

The horizon scan for repurposed medicines in development by non-commercial organisations revealed a total of 528 technologies [medicinal product (or combination of products) for a specified indication] with trial primary completion dates between 1 April 2020 and 31 March 2023. The identified clinical trials revealed the wide range of targeted indications, therapeutic areas, therapeutic classes, and classification of the repurposed technologies.

The remarkable increase in the number of clinical trials that started between 2017 and 2020 highlights the recent attention repurposing is gaining. The medicine repurposing approach was significantly adopted during the COVID-19 pandemic as a suitable option for the timely discovery of potential treatments for the novel infectious disease.^7^ Although clinical trials evaluating treatment options for COVID-19 were excluded from this scan, the recent increase in number of clinical trials on repurposed medicines shows the high acceptance of the concept to address treatment need.

It must be noted that 402 (76%) technologies were reported as solely sponsored by a non-industry organisation such as academia, hospitals, or charities. The lack of industry collaboration may pose legal, financial, or regulatory obstacles to the formal adoption or approval of these medicines in the new indications. As the proposed new indication may be outside the company’s business strategy, Fetro (2023) recommends an early initiation of an academia-industry collaboration to establish a ‘co-development framework’ where expectations and risks are effectively managed and mitigated.^16^ However, companies may intend to initiate a line extension for a new indication as part of their business strategy for patent extension, therefore collaboration with academia may be refused.^10,16^

The Innovation Observatory developed a novel evidence-based approach to classify innovative technologies, and this was employed to categorise the innovative medicines identified in the repurposed medicines scan.^15^ The classification for this scan includes repurposed technologies (combinations)[either on- or off-patent, or both]; off-patent/generic repurposed technologies; and branded/on-patent repurposed technologies. Most repurposed technologies in combination treated rare disease conditions (mostly rare cancers) by evaluating the efficacy of the synergistic effect of combination therapies. Some of the targeted rare cancers include acute myeloid leukaemia, diffuse large B-cell lymphoma, and pancreatic cancer. Similarly, most of the repurposed monotherapy technologies treated rare disease conditions, with an equal distribution among rare cancers and rare non-cancers. This is supported by findings by van den Berg S et al. (2021) which revealed that medicine repurposing for rare diseases is a significant area of research for non-commercial organisations such as academia and not-for-profit stakeholders.^17^ There are huge cost implications in the discovery and development of novel therapies for rare diseases, as only 5% of pharmaceutical companies are investing in this area. Repurposing, therefore, provides a faster and economical way to address this high unmet need.^9^

Repurposing represents a dynamic medicine development approach for disease areas with high mortality rates and limited treatment options. The scan revealed a diverse range of targeted therapeutic areas; with haematological cancers and lymphomas being the most frequently targeted. Haematological cancers have both high incidence and mortality rates, supporting the impetus for the development of more effective treatment options.^18^ The haematological cancers most targeted include acute myeloid leukaemia, chronic lymphocytic leukaemia, and multiple myeloma. The second most treated therapeutic area was gastrointestinal cancers, in which 54 technologies (10.2%) were evaluated as either monotherapies or combination therapies. Most cases of gastrointestinal cancers are diagnosed at an advanced stage where the prognosis is poor and survival rate greatly reduced.^19^ Chemotherapy, notably combination therapies, have been reported to improve patients’ survival and quality of life.^19,20^ This combination of high unmet need and potential for combination therapies to improve outcomes is likely driving the repurposing activity in this space, with the scan revealing that 36 of the 54 technologies (70.6%) treating gastrointestinal cancers were combination therapies. This trend of exploring combination therapies in areas of high unmet need merits further research. As more outcome data are collated, prediction of most promising combinations may become feasible.

A significant number of technologies treated neurological disorders (excluding neurological cancers) which was the third most targeted therapeutic area. The disorders targeted include amyotrophic lateral sclerosis (ALS), multiple sclerosis (MS), and Alzheimer’s disease. Treatment for neurological disorders requires a multi-disciplinary approach to reduce or manage symptoms to improve patients’ quality of life and reduce the global health burden.^21^ As there is currently no cure for ALS, MS or Alzheimer’s disease, researchers continue to explore different therapies to slow disease progression or manage symptoms.^22-24^

The therapeutic or disease areas least investigated in the scan, with six or fewer technologies, include: breast cancer; ear, nose, and throat; hepatology; pain relief; renal cancer; nephrology; skin cancer; male reproductive cancer; urological cancer; women’s and men’s health. Further work is required by industry and non-industry into the development of new treatment options for these therapeutic areas. There may have been off-label use of different medicinal products to treat some of these disease conditions, and these treatments may have been based on the clinicians’ discretion, however there is need for off-label benefits to be explored in pivotal clinical trials to potentially lead to licensing, and eventually, facilitate more equitable patient access.^13^

Monoclonal antibodies (such as pembrolizumab, obinutuzumab), kinase inhibitors (namely ibrutinib, carfilzomib), and corticosteroids (including dexamethasone, prednisone) were the top three drug classes being repurposed either as monotherapy or in combination with other medicinal products to treat the various therapeutic areas. The monoclonal antibodies targeted mostly cancers and immunological disorders. The diverse modes of action of monoclonal antibodies to directly target tumour cells while inducing anti-tumour immune response,^25^ make them promising candidates for these disease areas. The mechanism of action of kinase inhibitors to block the growth and survival of tumours,^26^ was explored as repurposed candidates to treat various cancers, most commonly haematological cancers and lymphomas. Corticosteroids (synthetic analogues of natural steroid hormones used for their anti-inflammatory, immunosuppressive and vasoconstrictive effects)^27^ were evaluated as monotherapies to treat cardiovascular or respiratory system disorders, while their use in various combinations with monoclonal antibodies and/or kinase inhibitors, was targeted at oncology indications.

Although every effort was made to ensure accuracy and completeness of the data, the manual, semi-automated and automated screening, and classification processes utilised in this study may mean some omissions are likely. Despite obvious limitations such as the specific inclusion criteria, these data show a rich and varied landscape of research in medicine repurposing, across a range of interventions, indications, and trial logistics.

## Conclusion

For a new medicine to be approved for use, regulatory authorities issue a Marketing Authorisation (MA) after a series of procedures to evaluate its safety and efficacy. For repurposed medicines, the MA holder is required to file a variation, line extension, or submit a new application for the new indication. However, in the case of repurposing by non-commercial organisations such as academia, support will be needed in the absence of a collaborating company. Additionally, a MA or product licence for a medicine does not necessarily mean patient access, hence further support is required to facilitate the adoption of the repurposed medicine into the health service for equitable patient access.

One purpose of the horizon scan for repurposed medicines is to support the identification of potential candidate medicines which will be assessed for suitability for the NHS England Medicines Repurposing Programme (MRP).^13^ Subsequent scans will provide systematic intelligence on future repurposing opportunities (an early alert process) which will allow the MRP ongoing early intelligence from which to assess and prioritise medicines that can benefit from tailored support towards licensing and adoption into the NHS. This support has the potential to overcome some of the barriers currently faced by non-commercial organisations. Support may also be provided through identifying a route for fast-track evidence generation and synthesis where evidence gaps are identified for prioritised medicine, facilitating licensing, or national policy or guidelines.^13^

This horizon scanning intelligence also offer audiences an opportunity to look strategically at activity and identify areas for collaboration and further research. Despite challenges which can prevent repurposing research and consequently delay patient access to effective medicines, there is potential for a systematic and transparent horizon scanning system to offer intelligence insights and support strategic research collaborations.^12^ The Innovation Observatory has expanded this work and integrated the identification, filtration, and prioritisation of repurposed medicines into its routine horizon scanning for innovative health technologies. This intelligence has been provided in an interactive living <u>dashboard</u> on the Innovation Observatory website as a source of intelligence on repurposed medicines, as well as to provide valuable insights into innovation trends, gaps, and areas of unmet medical need.^14^ This systematic approach will provide ongoing evidence on future repurposing opportunities of benefit to the NHS and the wider research ecosystem.

## Data Availability

All data produced in the present work are contained in the manuscript, and on the Repurposed medicines dashboard on the NIHR Innovation Observatory website https://www.io.nihr.ac.uk/dashboardpages/repurposed-medicines-dashboard/

https://www.io.nihr.ac.uk/dashboardpages/repurposed-medicines-dashboard/

## Authorship statement

SA, RF, AI, RP, and AO wrote the manuscript. SA, RF and DC conceived the design. RF, AI, AO, and RP contributed to data collection. SA and RP analysed and interpreted the data. All authors provided critical feedback and helped shape the research, analysis, and manuscript.

## Acknowledgments

We sincerely thank Amy Hussain, Hassina Carr, Lucy Imber, Jane Nesworthy, Olushola Ewedairo, Anjum Jahan and Sean Gill (NIHR Innovation Observatory) for their assistance in the collection of data for this project. We thank Dapo Ogunbayo for his contribution towards the delivery of the pilot scan; Dapo was employed by the NIHR Innovation Observatory during the early phase of this project.

## Reference

1 Morgan S, Grootendorst P, Lexchin J, Cunningham C, Greyson D. The cost of drug development: a systematic review. Health policy. 2011;100(1):4–17. 10.1016/j.healthpol.2010.12.002.

2 Hughes JP, Rees S, Kalindjian SB, Philpott KL. Principles of early drug discovery. British journal of pharmacology. 2011;162(6):1239–49. 10.1111%2Fj.1476-5381.2010.01127.x.

3 Pushpakom S, Iorio F, Eyers PA, Escott KJ, Hopper S, Wells A, et al. Drug repurposing: progress, challenges and recommendations. Nature reviews Drug discovery. 2019;18(1):41–58. 10.1038/nrd.2018.168.

4 Krayenbühl J, Vozeh S, Kondo-Oestreicher M, Dayer P. Drug–drug interactions of new active substances: mibefradil example. European journal of clinical pharmacology. 1999;55(8):559–65. 10.1007/s002280050673.

5 Kola I. The state of innovation in drug development. Clinical Pharmacology & Therapeutics. 2008;83(2):227–30. 10.1038/sj.clpt.6100479.

6 Breckenridge A, Jacob R. Overcoming the legal and regulatory barriers to drug repurposing. Nature reviews Drug discovery. 2019;18(1):1–2. 10.1038/nrd.2018.92.

7 Govender K, Chuturgoon A. An overview of repurposed drugs for potential COVID-19 treatment. Antibiotics. 2022;11(12):1678. 10.3390%2Fantibiotics11121678.

8 Jourdan J-P, Bureau R, Rochais C, Dallemagne P. Drug repositioning: a brief overview. Journal of Pharmacy and Pharmacology. 2020;72(9):1145–51. 10.1111%2Fjphp.13273.

9 Dhir N, Jain A, Mahendru D, Prakash A, Medhi B. Drug repurposing and orphan disease therapeutics. Drug Repurposing Hypothesis, Mol Asp Ther Appl. 2020. 10.5772/intechopen.91941.

10 Cha Y, Erez T, Reynolds I, Kumar D, Ross J, Koytiger G, et al. Drug repurposing from the perspective of pharmaceutical companies. British journal of pharmacology. 2018;175(2):168–80. 10.1111/bph.13798.

11 Konwar M, Bose D, Gogtay NJ, Thatte UM. Investigator-initiated studies: Challenges and solutions. Perspectives in clinical research. 2018;9(4):179. https://pubmed.ncbi.nlm.nih.gov/30319949/.

12 Del Álamo M, Bührer C, Fisher D, Griese M, Lingor P, Palladini G, et al. Identifying obstacles hindering the conduct of academic-sponsored trials for drug repurposing on rare-diseases: an analysis of six use cases. Trials. 2022;23(1):783. 10.1186/s13063-022-06713-y.

13 NHS England and NHS Improvement. Opportunities to repurpose medicines in the NHS in England. 2021. Available from: https://www.england.nhs.uk/wp-content/uploads/2021/03/B0342-oportunities-to-repurpose-medicines-in-the-nhs-in-england.pdf [Accessed 24th March 2023].

14 NIHR Innovation Observatory (IO). Repurposed medicines dashboard. 2024. Available from: https://www.io.nihr.ac.uk/dashboardpages/repurposed-medicines-dashboard/ [Accessed 5th March 2024].

15 Ogunbayo D, Coughlan D, Fairbairn R, Akinbolade S. Classification System for Innovative Medicines in the Pipeline: New or Repurposed? HTAi 2022. 2022. 10.1017/s0266462322001088.

16 Fetro C. Connecting academia and industry for innovative drug repurposing in rare diseases: it is worth a try. Rare Dis. Orphan Drugs J. 2023;2(7). 10.20517/rdodj.2023.06.

17 van den Berg S, de Visser S, Leufkens HG, Hollak CE. Drug repurposing for rare diseases: a role for academia. Frontiers in Pharmacology. 2021;12:746987. 10.3389/fphar.2021.746987.

18 Michael AA, Balakrishnan P, Velusamy T. Drug Repurposing for Hematological Malignancies. In: Drug Repurposing for Emerging Infectious Diseases and Cancer. Springer; 2023: 217–52.

19 Araújo D, Ribeiro E, Amorim I, Vale N. Repurposed drugs in gastric cancer. Molecules. 2022;28(1):319. 10.3390/molecules28010319.

20 Wagner AD, Syn NL, Moehler M, Grothe W, Yong WP, Tai BC, et al. Chemotherapy for advanced gastric cancer. Cochrane database of systematic reviews. 2017;(8). 10.1002/14651858.CD004064.pub4.

21 National Health Service (NHS) Inform. Brain, nerves and spinal cord. 2023. Available from: https://www.nhsinform.scot/illnesses-and-conditions/brain-nerves-and-spinal-cord/ [Accessed 22nd March 2024].

22 National Institute of Neurological Disorders and Stroke. Amyotrophic Lateral Sclerosis (ALS). 2024. Available from: https://www.ninds.nih.gov/health-information/disorders/amyotrophic-lateral-sclerosis-als#toc-how-is-amyotrophic-lateral-sclerosis-als-diagnosed-and-treated- [Accessed15th March 2024].

23 Alzheimer’s Research UK. Treatments for Alzheimer’s. 2022. Available from: https://www.alzheimersresearchuk.org/dementia-information/types-of-dementia/alzheimers-disease/treatments/ [Accessed 15th March 2024].

24 National Health Service (NHS). Multiple sclerosis: Treatment. 2022. Available from: https://www.nhs.uk/conditions/multiple-sclerosis/treatment/ [Accessed 19th April 2024].

25 Zahavi D, Weiner L. Monoclonal antibodies in cancer therapy. Antibodies. 2020;9(3):34. 10.3390%2Fantib9030034.

26 National Cancer Institute. Kinase inhibitors. 2024. Available from: https://www.cancer.gov/publications/dictionaries/cancer-terms/def/kinase-inhibitor [Accessed 15th March 2024].

27 National Library of Medicine. Corticosteroids. 2023. Available from: https://www.ncbi.nlm.nih.gov/books/NBK554612/ [Accessed 15th March 2024].

